# Potential protective link between type I diabetes and Parkinson’s disease risk and progression

**DOI:** 10.1101/2022.10.12.22280972

**Authors:** Konstantin Senkevich, Paria Alipour, Ekaterina Chernyavskaya, Eric Yu, Alastair J Noyce, Ziv Gan-Or

## Abstract

**Background:** Epidemiological studies suggested an association between Parkinson’s disease (PD) and type 2 diabetes, but less is known about type 1 diabetes (T1D) and PD.

**Objectives:** To explore the association between T1D and PD.

**Methods:** We used Mendelian randomization, linkage disequilibrium score regression and transcriptome wide association analysis (TWAS) to examine the association between PD and T1D.

**Results:** Mendelian randomization showed a potentially protective role of T1D for PD risk (inverse-variance weighted (IVW); OR (95% CI) 0.97 (0.94-0.99); p=0.039), as well as motor (IVW; 0.94 (0.88-0.99); p=0.044) and cognitive progression (IVW; 1.50 (1.08-2.09); p=0.015). We further found negative genetic correlation between T1D and PD (rg=-0.17, p=0.016), and identified nine genes in cross-tissue TWAS that were associated with both traits.

**Conclusions:** Our results suggest a potential genetic link between T1D and PD risk and progression. Larger comprehensive epidemiological and genetic studies are required to validate our findings.

## Introduction

Multiple lines of evidence suggest an association between type 2 diabetes (T2D) and Parkinson’s disease (PD).^1-4^ T2D is associated with both increased PD risk and worse progression, measured by cognitive and motor scales.^4^ Moreover, drugs targeting TD may reduce the risk of PD and potentially could be repurposed to modify PD progression.^5^

Less is known about the link between PD and type 1 diabetes (T1D). T1D is an autoimmune disorder characterized by the destruction of islets of Langerhans in the pancreas.^6^ The pathophysiology of T1D is different from T2D; nonetheless both diseases have strong genetic correlation and shared biological pathways.^7^ PD is a complex disease with multiple pathways involved in its development,^8^ including pathways related to immune response and inflammation.^9^ Most observational studies did not differentiate between T1D and T2D when defining diabetes as a risk factor,^10-12^ since T1D is much less prevalent than T2D. One report suggested a potential increased risk of PD in patients with T1D.^13^

Mendelian randomization (MR) uses genetic variants such as single nucleotide polymorphisms (SNPs) associated with an exposure of interest (in our case, T1D) as proxies for causal inference about the association between that exposure and an outcome. In the current study we performed MR to estimate whether a relationship between T1D and PD risk and progression may exist. Furthermore, we conducted genetic correlation analysis and transcriptome-wide association study (TWAS) to assess potential shared genetic architecture.

## Methods

### Mendelian Randomization

We selected publicly availably genome-wide association studies (GWASs) for T1D and PD risk and progression with participants of European ancestry and no overlapping samples. We used SNPs from the selected GWASs that were significant at GWAS level (p<5×10^−8^) to construct a genetic instrument for the exposure (T1D) and examine its effects on two categories of outcomes: PD risk and PD progression. For the exposure we selected a recent T1D study (N cases= 13,458; N controls= 20,143) ^14^ downloaded from the GWAS catalog,^15^ with only samples of European ancestry being included. For the outcome, we selected the most recent PD GWAS (N cases= 33,674; N controls= 449,056).^16^ UK biobank participants were included in the PD GWAS but not in the selected T1D study, to avoid potential bias.

To study the genetically estimated effect of T1D on PD motor progression, measured by Unified Parkinson Disease Rating Scale (UPDRS) Part III, and on PD cognitive progression, measured by Montreal Cognitive Assessment (MoCa) and Mini-Mental State Exam (MMSE), we selected the largest publicly available GWASs of these continuous traits.^17^ The GWAS on PD progression is a meta-analysis of several studies, with different number of participants for each phenotype. It means that results for different SNPs correspond for different number of cases. Therefore, to calculate the sample sizes for PD progression studies, we calculated the means of patients included in each analysis across all SNPs. The mean sample sizes included in the GWASs of PD progression traits were N=1,398 for UPDRS Part III, N=1,329 for MMSE and N=1,000 for MoCA.

To perform MR we used the Two-sample MR R package.^18, 19^ We applied Steiger filtering, to exclude SNPs that explain more variance in the outcome than in the exposure.^19^ We used inverse variance weighted (IVW) meta-analysis, which combines results from individual Wald ratios together. We used MR Egger, which likewise combines separate Wald ratios into meta-regression to obtain an estimate that is unbiased in the presence of directional pleiotropy.^20^ Considering that some of our IVs could be invalid, we also used weighted median based estimate to account for it.^21^ To further explore potential pleiotropy, a variety of sensitivity analysis were applied including Cochran’s Q test in the IVW, MR-Egger methods and global MR-PRESSO.^22^ We calculated power to detect an equivalent effect size of OR 1.2 on PD risk and progression utilizing an online Mendelian randomization power calculation https://sb452.shinyapps.io/power/).^23^

### Genetic correlation

We examined genetic correlations between PD and T1D using linkage disequilibrium score regression (LDSC) as previously described.^24, 25^ LDSC considers linkage disequilibrium structure to estimate potential genetic overlap between two traits. MR analyses and genetic correlation were done after the exclusion of SNPs within the major histocompatibility complex (MHC) region due to the biased linkage disequilibrium structure.

### Transcriptome wide association analysis

To calculate cross-tissue, gene-expression associations in T1D and PD we used the Unified Test for Molecular Signatures (UTMOST) software.^26^ We used a pre-calculated matrix with tissue-specific TWAS weights, which was created using grouped penalized regression. In the next step, we used UTMOST to conduct single-tissue TWASs across 44 tissues available in GTEx (V6). Subsequently, we used UTMOST to define genes associated with T1D and PD across all tissues, by combining the single-tissue test results with Generalized Berk-Jones (GBJ) test.^26^ Finally, we applied false discovery rate correction and did head-to-head comparisons of genes significant for both PD and T1D.

### Data availability

We used only publicly available data in the current study. References for GWASs and packages for analysis are detailed in the Methods section. All results are reported in the tables or attached in the supplementary data.

## Results

### Evidence for a modest protective effect of T1D on PD risk and progression

The instruments in all analyses had sufficient strength as demonstrated by F-statistics >10 (Table 1). We found weak evidence of a modest protective effect of T1D on PD risk (IVW; OR=0.97, 95% CI 0.94-0.99, p=0.039; weighted median; OR=0.95, 95% CI 0.90-0.99, p=0.026, Table 1, Supplementary figure 1A). We studied the effects of T1D on motor and cognitive progression. UPDRS3 is a motor performance scale, meaning that higher scores indicate poorer performance. MMSE and MoCA are cognitive scales, and higher scores signify better performance. We observed potentially protective effects of T1D on motor progression measured by UPDRS3 (IVW, OR=0.94, 95% CI 0.88-0.99, p=0.044, Table 1, Supplementary figure 1B) and on cognitive progression as measured by both MMSE (IVW, OR=1.11, 95% CI 0.99-1.25, p=0.060) and MoCA (IVW, OR=1.50, 95% CI 1.08-2.09, p=0.015, Table 1, Supplementary figure 1C-D). We found that rs7110099 near *INS-IGF2* (Insulin-Insulin like growth factor 2) and rs56994090 near *MEG3* (Maternally expressed gene 3) have potential protective effects on PD risk (Wald ratio OR=0.95, 95% CI 0.89-1.00, p=0.055 and OR=0.67, 95% CI 0.46-0.96, p=0.03, respectively, uncorrected p values). Another SNP next to *MEG3*, rs4900384, might have a protective effect for cognitive progression as measured by MoCA (Wald ratio OR=17.25, 95% CI 1.86-159.50, p=0.010). We did not find pleiotropy in any of sensitivity analysis (Supplementary Table 1). Furthermore, we applied MR-PRESSO analysis and did not find either general pleiotropy or specific pleiotropic SNPs (Supplementary Table 1).

**Table 1.** MR analysis between exposure T1D and outcome PD risk and progression.

| Outcome | N, SNPs included | Power, % | F-statistics | Inverse variance weighted |  | MR Egger |  |
| --- | --- | --- | --- | --- | --- | --- | --- |
|  |  |  |  | OR (95% CI) | P | OR (95% CI) | P |
| **PD risk** | 58 | 100 | 22 | 0.97 (0.94-0.99) | **0.039** | 0.96 (0.91-1.01) | 0.115 |
| **UPDRS 3** | 42 | 32.8 | 34 | 0.94 (0.88-0.99) | **0.044** | 1.03 (0.88-1.20) | 0.721 |
| **MMSE** | 39 | 31.4 | 37 | 1.11 (0.99-1.25) | 0.060 | 1.13 (0.85-1.50) | 0.418 |
| **MoCA** | 38 | 24.8 | 39 | 1.50 (1.08-2.09) | **0.015** | 2.00 (0.85-4.72) | 0.121 |

PD, Parkinson's disease; T1D, type 1 diabetes; P, P-value; MR, Mendelian randomization; UPDRS3, unified Parkinson's disease rating scale part 3; MMSE, Mini Mental State Examination; MoCA, Montreal Cognitive Assessment; OR, odds ratio; 95%CI- 95% confidence interval.

### Shared expression for genes related to autophagy and lysosomal pathways between T1D and PD

We found evidence for some negative genetic correlation between T1D and PD using LDSC (rg=-0.17, p=0.016). We then performed TWASs on T1D and PD in multiple tissues and selected significant genes across all tissues for both traits after FDR correction. We demonstrated nine significant genes for PD as well as for T1D (Table 2, *AAR2, CTSB, LAT, LRRC37A, LRRC37A2, R3HDM1, RAB7L1, RNF40, WNT3*) suggesting potential pleiotropy that was not detected by the MR tools.

**Table 2.** Genes associated with both T1D and PD in cross-tissue transcriptomic gene-trait association analysis.

| Gene | T1D | PD |
| --- | --- | --- |
|  | Pfdr | Pfdr |
| *AAR2* | 0.030 | 0.041 |
| *CTSB* | 8.06E-05 | 0.028 |
| *LAT* | 0.026 | 0.009 |
| *LRRC37A* | 5.25E-04 | 0.0001 |
| *LRRC37A2* | 8.21E-04 | 4.36E-08 |
| *R3HDM1* | 0.046 | 0.001 |
| *RAB7L1* | 0.001 | 1.65E-05 |
| *RNF40* | 0.027 | 0.002 |
| *WNT3* | 0.010 | 2.24E-08 |

Pfdr, P-value after false discovery rate correction; T1D, type 1 diabetes; PD, Parkinson's disease

## Discussion

In this analysis, we demonstrated a potential protective effect of T1D on PD risk and progression. We did not find obvious pleiotropy or heterogeneity in any of our MR analyses, using a variety of sensitivity methods. However, our genetic correlation analysis suggested that there is some potential pleiotropy, with negative genetic correlation between the two traits (i.e. variants that are associated with reduced risk of one trait are associated with increased risk of the other traits). This may suggest that the association seen in the MR analysis is due to residual pleiotropy that was not identified by the MR tools. This observation was reinforced by our cross-tissue TWASs, demonstrating that several genes may be overlapping between the two traits.

We demonstrated potential protective effects of SNPs near *IGF2* and *MEG3* for PD. Previously, neuroprotective effect of *IGF2* was reported in cell and mouse models of PD ^27^ and its downregulation was shown in PD patients’ blood.^28^ Moreover, overexpression of *IGF2* resulted in neuroprotective effect.^29^ Similarly, downregulation of *MEG3* was recently reported in PD patients ^30^ and its overexpression could be protective for PD through negative regulation of LRRK2.^30^

Inflammatory and autoimmune pathways play an important role in the development of T1D.^31^ Accumulating evidence suggest lysosomal dysfunction as a prevalent mechanism in the pathogenesis of PD.^32^ We showed that *CTSB* and *RAB7L1* were associated in cross-tissue TWAS analysis with both PD and T1D. These genes are playing an important role in the autophagy-lysosome pathway, suggesting a role for lysosomal function in both traits and potential pathway overlap. Another gene in overlap between these traits was *CD19*, which is encoding B-lymphocyte antigen CD19, demonstrating the potential importance of immune pathways for both traits outside of the MHC locus. Previously, using the conjunction false discovery rate method, some weak pleiotropy was demonstrated between PD and T1D,^33^ further supported by our findings. Recently, similar protective effect in MR study was demonstrated for another autoimmune disease – rheumatoid arthritis.^34^ The authors also highlighted the hypothesis that the protective effect of autoimmune conditions of PD could be driven by some variants in genes involved the lysosomal-autophagy pathway.^34^ We suggest that protective association could be driven by pleiotropy, particularly in lysosomal genes, as demonstrated by the genetic correlation and TWAS in our analyses.

Our study has several limitations. First, we only included samples of European ancestry, since large GWASs in other populations do not exist, therefore our findings cannot be generalized to the population at large. Second, the GWASs on PD progression parameters are underpowered (<80%). Thus, additional replication is required whenever larger GWASs on PD progression is available. Lastly, MR could be influenced by the quality of selected GWASs that are used for the MR analysis. We therefore used different GWASs for exposure and non-overlapping cohorts in the outcome to partially account for this limitation.

To conclude, our results support a protective effect of T1D on PD risk and progression, that could be driven by potential pleiotropy. Larger comprehensive epidemiological studies are required to support further explore this association.

## Data Availability

All results are reported in the tables or attached in the supplementary data.

## Acknowledgment

ZGO is supported by the Fonds de recherche du Québec - Santé (FRQS) Chercheurs-boursiers award, and is a William Dawson Scholar and a Killam Scholar. KS is supported by a postdoctoral fellowship from the Canada First Research Excellence Fund (CFREF), awarded to McGill University for the Healthy Brains for Healthy Lives initiative (HBHL) and postdoctoral fellowship from Fonds de recherche du Québec - Santé (FRQS).

## Authors’ Roles

1. Research project: A. Conception, B. Organization, C. Execution;
2. Statistical Analysis: A. Design, B. Execution, C. Review and Critique;
3. Manuscript Preparation: A. Writing of the first draft, B. Review and Critique.

KS: 1A, 1B, 1C, 2A, 2B, 3A

PA: 1C, 3B

EC: 1C, 3B

EY: 1C, 2C, 3B

AJN: 1A, 2C, 3B

ZGO: 1A, 1B, 2C, 3B

## Financial Disclosures of all authors (for the preceding 12 months)

ZGO received consultancy fees from Lysosomal Therapeutics Inc. (LTI), Idorsia, Prevail Therapeutics, Inceptions Sciences (now Ventus), Ono Therapeutics, Denali, Handl Therapeutics, Neuron23, Bial Biotech, UCB, Guidepoint, Lighthouse and Deerfield. Other authors have nothing to disclose.

