## Supplementary Table 1 for "Potential protective link between type I diabetes and Parkinson’s disease risk and progression"

Supplementary Table 1. MR sensitivity analysis between exposure T1D and outcome PD risk and progression.

| **Outcome** | **Heterogeneity tests** | | | | | | **Test for directional horizontal pleiotropy** | | | |
| --- | --- | --- | --- | --- | --- | --- | --- | --- | --- | --- |
|  | **MR Egger** | | | **Inverse variance weighted** | | |  |  |  |  |
|  | **Q** | **Q_df** | **Q_pval** | **Q** | **Q_df** | **Q_pval** | **Egger intercept** | **se** | **pval** | **MR-PRESSO global (pval)** |
| **PD risk** | 53.5 | 56 | 0.571 | 53.730 | 57 | 0.599 | 0.002 | 0.005 | 0.614 | 0.554 |
| **UPDRS3** | 28.1 | 40 | 0.920 | 29.680 | 41 | 0.910 | -0.017 | 0.012 | 0.216 | 0.909 |
| **MMSE** | 37.5 | 37 | 0.446 | 37.507 | 38 | 0.492 | -0.002 | 0.022 | 0.935 | 0.360 |
| **MoCA** | 41.8 | 36 | 0.233 | 42.390 | 37 | 0.250 | -0.049 | 0.068 | 0.481 | 0.271 |

Q- Cochran’s Q test, df- degrees of freedom, se- standard error, T1D- type 1 diabetes; UPDRS3- Unified Parkinson Disease Rating Scale Part III and MoCA- Montreal Cognitive Assessment, MMSE- Mini-Mental State Exam
