## Supplementary Figure 1 for "Potential protective link between type I diabetes and Parkinson’s disease risk and progression"

Supplementary Figure 1 A-D. Forest plots showing point estimates of the Type 1 diabetes risk on outcome of interest, outcome of interest at the top of each forest plot.

Black points represent log-odds ratio of each SNP on the risk of PD. Red points represent the log-odds ration when combining all SNPs together (Inverse variance weighted and MR Egger methods). Lines from points represent 95% confidence intervals.

**Supplementary Figure 1A Parkinson’s disease risk as outcome**
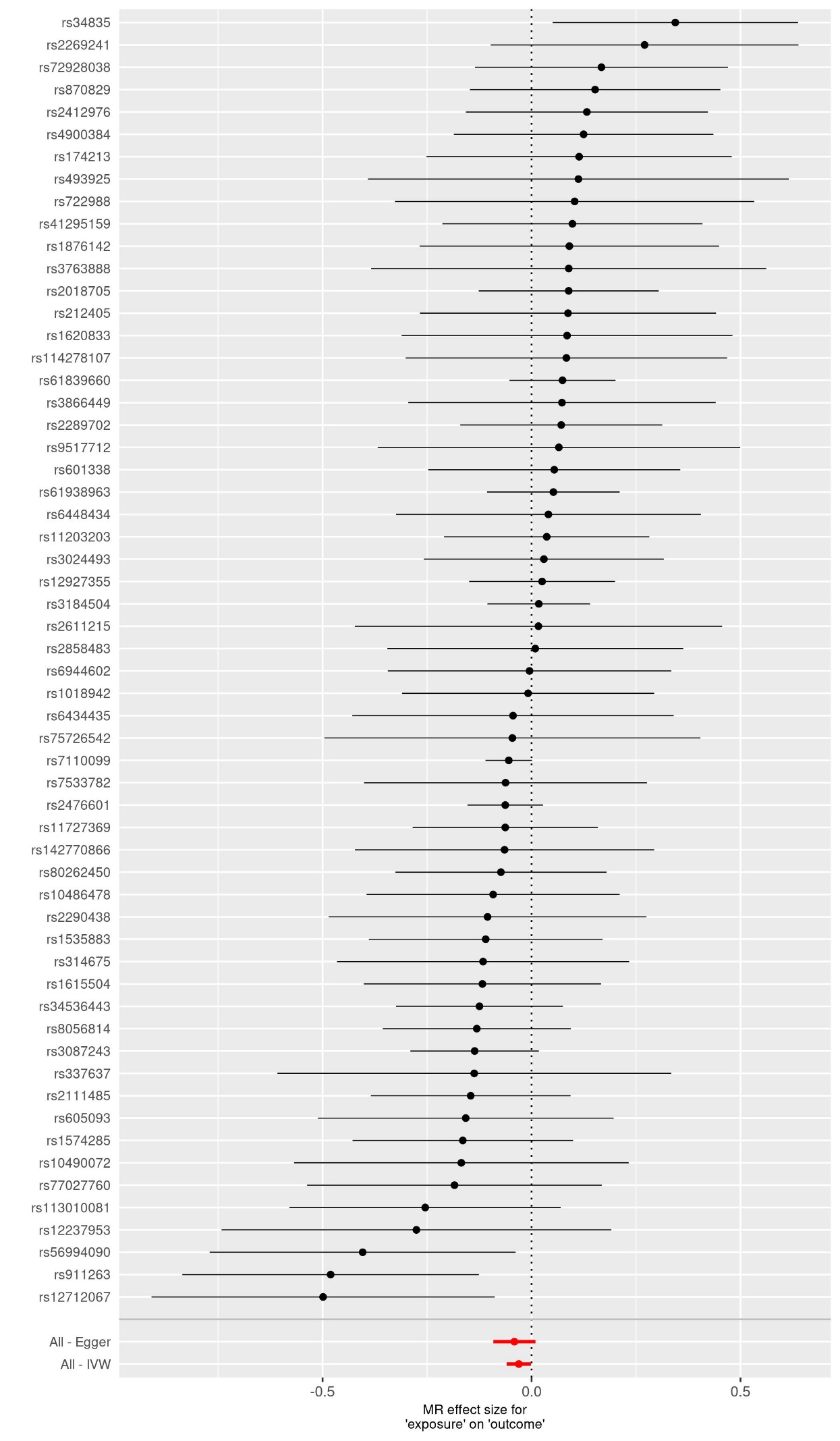

**Supplementary Figure 1B UPDRS3 as outcome**
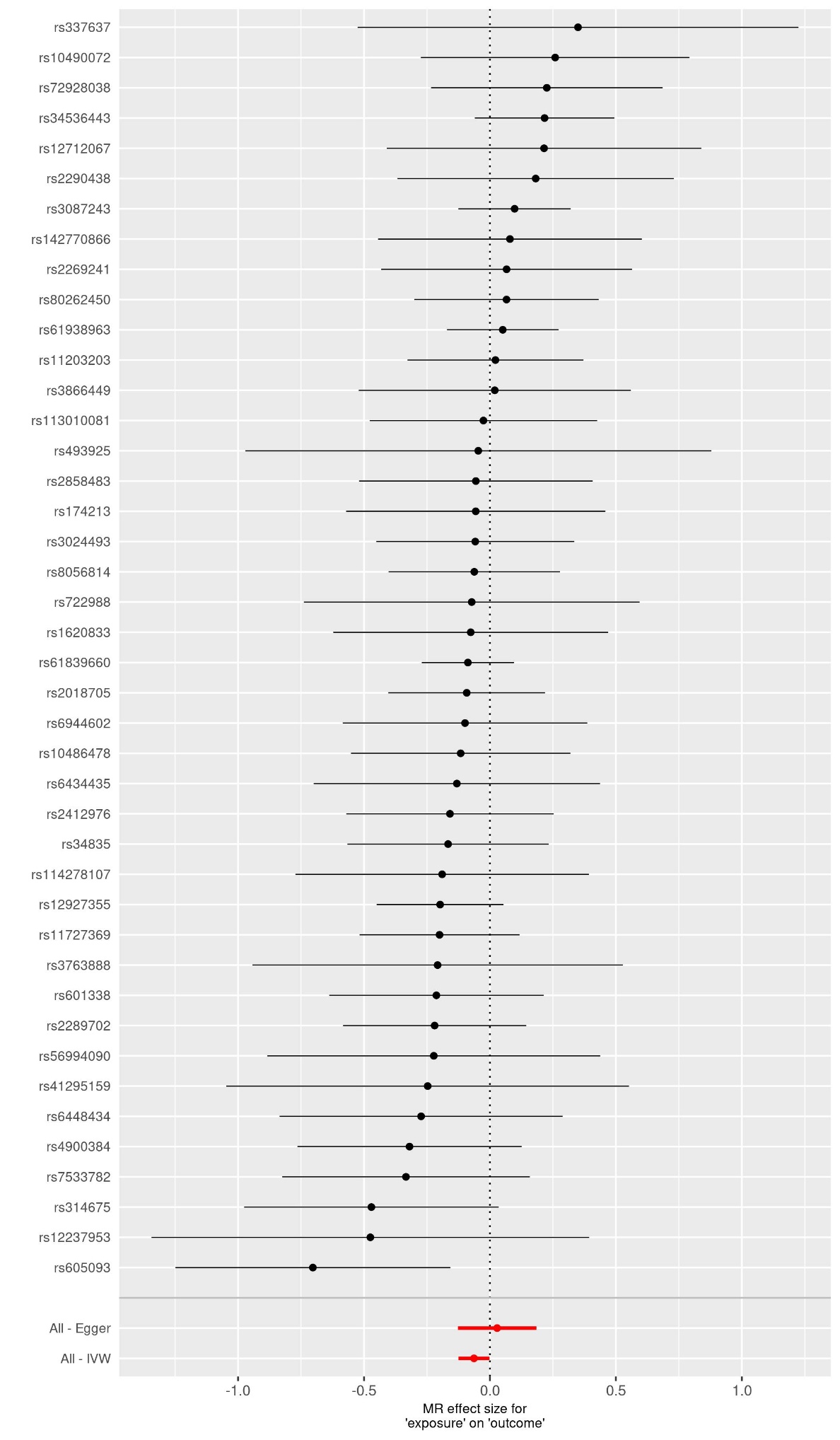

**Supplementary Figure 1C MoCA as outcome**
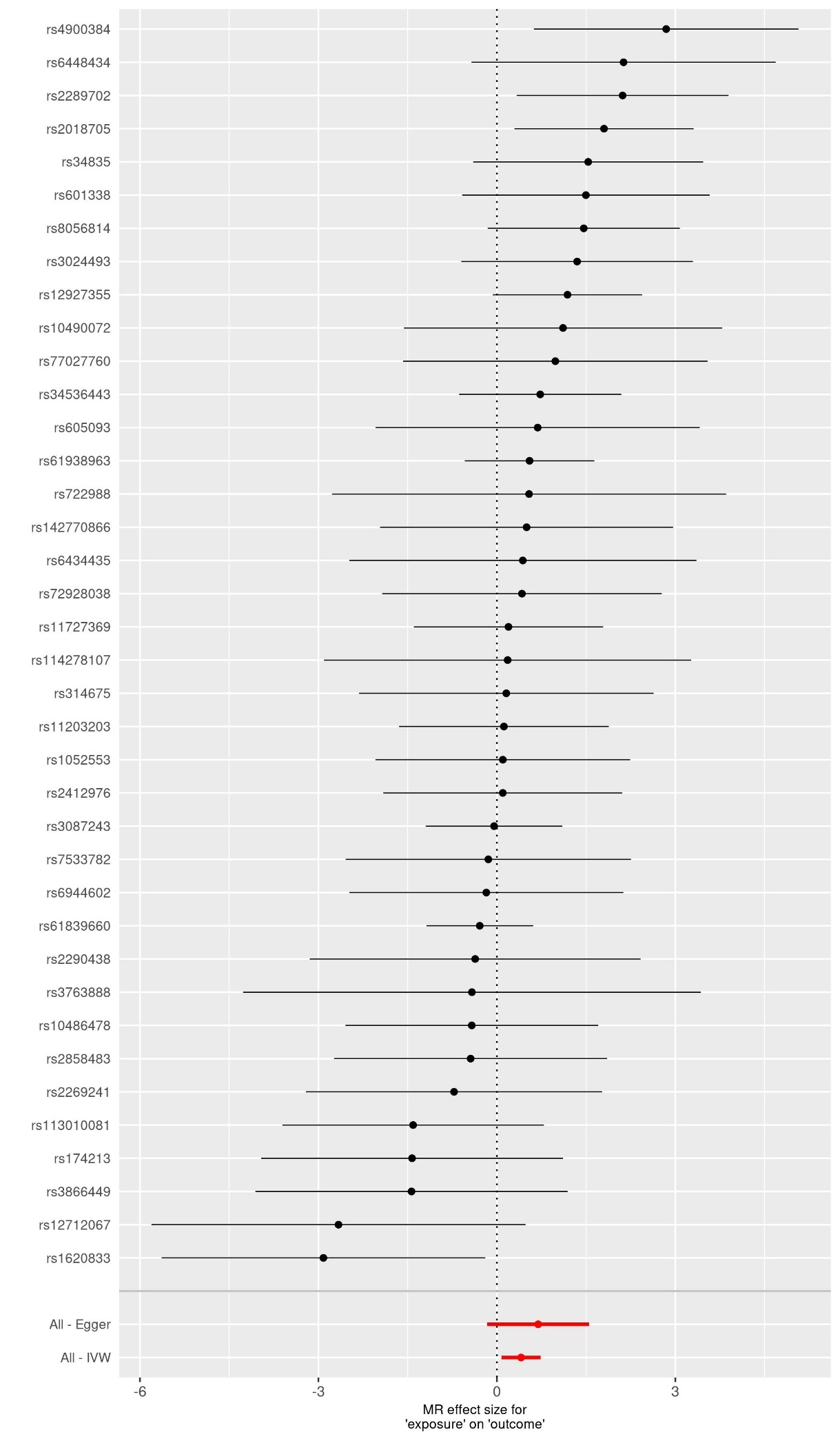

**Supplementary Figure 1D MMSE as outcome**

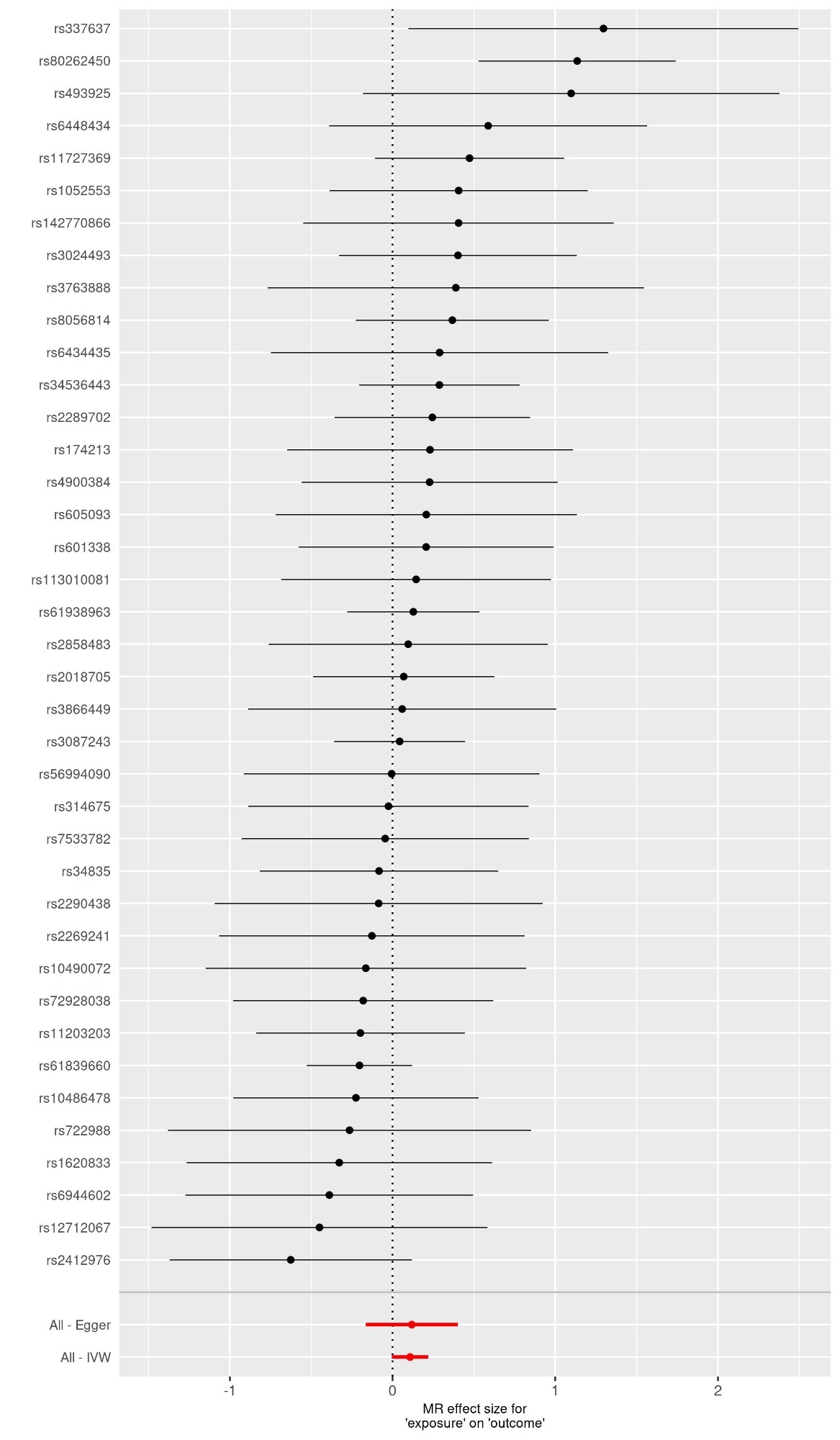
